# Antibody kinetics to SARS-CoV-2 at 13.5 months, by disease severity

**DOI:** 10.1101/2021.09.10.21262527

**Authors:** Concepción Violán, Pere Torán, Bibiana Quirant, Noemi Lamonja-Vicente, Lucía A. Carrasco-Ribelles, Carla Chacón, Josep Maria Manresa-Dominguez, Francesc Ramos-Roure, Albert Roso-Llorach, Aleix Pujol, Dan Ouchi, Mónica Monteagudo, Pilar Montero, Rosa Garcia-Sierra, Fernando Arméstar, Rosalia Dacosta-Aguayo, Maria Doladé, Nuria Prat, Josep Maria Bonet, Bonaventura Clotet, Ignacio Blanco, Julia G Prado, Eva María Martínez Cáceres, for the ProHEpiC-19 Investigators

**Affiliations:** Unitat de Suport a la Recerca Metropolitana Nord, Institut Universitari d’Investigació en Atenció Primària Jordi Gol (IDIAP Jordi Gol), Mataró, Spain; Direcció d’Atenció Primària Metropolitana Nord Institut Català de Salut; Germans Trias i Pujol Research Institute (IGTP), Badalona, Spain; Universitat Autònoma de Barcelona, Cerdanyola del Vallès, Spain; Immunology Department. FOCIS Center of Excellence- Universitat Autònoma de Barcelona, Cerdanyola del Vallès, Spain; Immunology Division. Laboratori clinic Metropolitana Nord (LCMN). Hospital Universitari Germans Trias i Pujol, Badalona, Spain; Institut Universitari d’Investigació en Atenció Primària Jordi Gol (IDIAP Jordi Gol), Barcelona, Spain; Department of Nursing, Universitat Autònoma de Barcelona, Cerdanyola del Vallès, Spain; Department of Medicine, Faculty of Medicine, Universitat Autónoma de Barcelona, 08193 Bellaterra, Spain; Departament de Pediatria, d’Obstetrícia i Ginecologia i de Medicina Preventiva, Universitat Autónoma de Barcelona, 08193 Bellaterra, Spain; AIDS Research Institute Irsicaixa, Badalona, Spain; Grup de REcerca Multidisciplinar en SAlut i Societat (GREMSAS), (2017 SGR 917), 08007 Barcelona, Spain; Intensive Care Unit, Hospital Universitari Germans Trias i Pujol, Badalona, Spain; Clinical and Biochemical Analysis Division. Laboratori clinic Metropolitana Nord (LCMN). Hospital Universitari Germans Trias i Pujol, Badalona, Spain; Lluita contra la SIDA Foundation, Hospital Universitari Germans Trias i Pujol, Badalona, Spain; Hospital Universitari Germans Trias i Pujol, Badalona, Spain; Gerència Territorial Metropolitana Nord, Institut Català de la Salut, Barcelona, Spain

**Keywords:** COVID-19, Antibodies, IgG, IgM, Seroprevalence, Kinetics, Humoral immunity, Disease spectrum, Sex, Health Care Workers

## Abstract

**Background:** Understanding humoral responses and seroprevalence in SARS-CoV-2 infection is essential for guiding vaccination strategies in both infected and uninfected individuals.

**Methods:** We determine the kinetics of IgM against the nucleocapsid (N) and IgG against the spike (S) and N proteins of SARS-CoV-2 in a cohort of 860 health professionals (healthy and infected) in northern Barcelona. We model the kinetics of IgG and IgM at nine time points over 13.5 months from infection, using non-linear mixed models by sex and clinical disease severity.

**Results:** Of the 781 participants who were followed up, 478 (61.2%) became infected with SARS-CoV-2. Significant differences were found for the three antibodies by disease severity and sex. At day 270 after diagnosis, median IgM(N) levels were already below the positivity threshold in patients with asymptomatic and mild-moderate disease, while IgG(N, S) levels remained positive to days 360 and 270, respectively. Kinetic modelling showed a general rise in both IgM(N) and IgG(N) levels up to day 30, followed by a decay whose rate depended on disease severity. IgG(S) levels increased at day 15 and remained relatively constant over time.

**Conclusions:** We describe kinetic models of IgM(N) and IgG(N, S) SARS-CoV-2 antibodies at 13.5 months from infection and disease spectrum. Our analyses delineate differences in the kinetics of IgM and IgG over a year and differences in the levels of IgM and IgG as early as 15 days from symptoms onset in severe cases. These results can inform public health policies around vaccination criteria.

Funded by the regional Ministry of Health of the Generalitat de Catalunya (Call COVID19-PoC SLT16_04; NCT04885478)

## BACKGROUND

Infection with SARS-CoV-2 can be determined by measuring the generation of virus-specific antibodies, reflecting an immune response against a recent or previous infection ^1^. Different studies described the rapid response of immunoglobulins (IgS) of various isotypes (IgA, IgM, IgG) against epitopes of the spike (S) glycoprotein as well as to the nucleocapsid (N) protein in SARS-CoV-2 infection ^2–5^.

COVID-19 has a broad clinical spectrum, with patients experiencing everything from asymptomatic infections to critical illness. Likewise, the antibody response to SARS-CoV-2 infection is heterogeneous ^6^. Few longitudinal studies have performed serological follow-up across the clinical spectrum, and with limited study periods, from 80 to 270 days ^2–4^. An early study reported a rapid rise and subsequent fall of antibodies, which stabilized at later time points, indicating that immunity against SARS-CoV-2 lasted for at least four months after infection ^7^. Two later studies extended this protection up to at least six months ^8, 2^, while recent estimates venture that it persists for at least a year ^9, 10^. However, knowledge of the kinetics and nature of the immune response to SARS-CoV-2 infection is still limited, and larger longitudinal studies are needed to define the half-life of antibodies against SARS-CoV-2 and their respective kinetics. Antibodies developed against the virus are directed to different regions of the spike (S) protein and/or against the nucleocapsid (N) or envelope proteins of SARS-CoV-2 ^11^. The vaccines are based only in the generation of antibodies against the S protein ^12^.

Available reagents in compliance with WHO International Standards for anti-SARS-CoV-2 antibodies allow us to evaluate vaccine efficacy and compare epidemiological and immunological surveillance studies. At the beginning of the pandemic, antibody levels were determined using qualitative assays; the availability of an internationally standardized enzyme immunoassay for the quantitative detection of specific IgG antibodies against the spike protein is recent ^13^.

The kinetics of the serological response to SARS-CoV-2 along the clinical spectrum will be key to define revaccination criteria, and guide public health policies. To achieve this, it is essential to know the half-life of antibodies against SARS-CoV-2 and their relationship with disease severity. The aim of this study was to describe the kinetics of IgM (N) and IgG (N, S) antibodies against SARS-CoV-2 and to assess the relationship between the immune response and the COVID-19 clinical spectrum.

## METHODS

### ProHEpiC-19 study design and cohort procedures

*ProHEpiC-19 is* a prospective, dynamic longitudinal study, involving two cohorts of health professionals (healthy and infected) in northern metropolitan Barcelona (Spain). The ethics committees of the IDIAPJGol Foundation (ref. 20/067) and IGTP Health Institute (ref. COV20/00660 (PI-20-205)) approved the study protocol (supplementary protocol, NCT04885478).

Health professionals were recruited from 3 March 2020 to 22 March 2021. Participants were allocated to their cohort (infected or uninfected) based on the following inclusion criteria: for non-infected participants, a negative serological test for IgM(N) and IgG(N), and negative RT-PCR at baseline; and for infected participants, confirmed COVID-19 by RT-PCR or antibodies against SARS-CoV-2. Exclusion criteria were refusal to participate or unavailable for follow-up. The first follow-up was on 5 May 2020 and the last on 14 May 2021.

The analysis reported in this work includes only participants with SARS-CoV-2 antibodies. Briefly, COVID-19-specific symptoms were recorded during the baseline clinical visit, and an RT-PCR test with nasal and oropharyngeal swab was performed and repeated at week one. In addition, serological tests were repeated at 15, 30, 60, 90, 180, 270, and 360 days following the baseline visit. Infected participants were classified into three different groups according to their symptomology: 1) asymptomatic: no symptoms; 2) mild-moderate: people with one or more clinical symptoms characteristic of COVID -19 who did not require hospital admission; 3) severe-critical: patients who required hospital and/or ICU admission.

### SARS-CoV-2 detection and quantification of IgM and IgG

RT-PCR was used as a diagnostic test. RNA for RT-PCR testing was extracted from fresh samples using the STARMag 2019-nCoV kit on a liquid-dispensing robot, and RNA detection was performed using the Allplex SARS-CoV-2 assay, a multiplex RT PCR assay to detect four SARS-CoV-2 target genes in a single tube. In addition, we conducted a pre-validation study (with six different IVD-CE-approved ELISA tests) and selected commercially available anti-SARS-CoV-2 IgG and IgM anti-N ELISA kits based on their performance. Participants with positive anti-N serology or/and RT-PCR were also tested for antibodies against the spike (S) subunit of SARS-CoV-2 by means of an enzyme immunoassay (ELISA) for the quantitative determination of IgG class antibodies using DECOV1901.

### Sample size

Sample size calculation for healthy and infected cohorts can be found in the supplementary protocol. The total collected sample of 478 infected participants achieves 100% power to detect differences among the means versus the alternative of equal means using an F test, assuming values of α=0.05 and an effect size of η^2^ =0.06. This calculation was carried out using a one-way ANOVA test with sample sizes of 72, 367, and 39 from the three clinical groups whose means were to be compared.

### Statistical analysis

A descriptive analysis was undertaken for all categorical and continuous variables. Missing values were found only in in the following sociodemographic variables: education (3.5%), marital status (4.6%), and nationality (7.5%). For the analysis of serological test results, only available data were used.

The evolution of antibody test results from diagnosis was studied considering both dichotomous (i.e. positive/negative result) and numerical values. For the dichotomous response, a descriptive analysis was performed to study the number of participants with each response pattern over time. The monitoring of antibody values after diagnosis was studied in three ways. First, we stratified antibody levels by days since diagnosis, describing them by boxplots and comparing them using statistical tests. Then, at each timepoint, differences were assessed according to disease severity (Holm-adjusted Dunn’s test) and sex (Mann-Whitney U test). Second, we fitted locally estimated scatterplot smoothing models (LOESS), calculating their associated 95% confidence intervals (CIs). Finally, non-linear mixed effects models (NLME) were also fitted. In these models, each parameter was assumed to have a fixed and a random effect. Both LOESS and NLME models were first fitted for all patients and then stratified by clinical condition and sex. The estimated NLME curves were used to model IgM and IgG kinetics over time. Model diagnostics were performed with residual analysis, and goodness-of-fit was checked with Akaike and Bayesian information criteria. For these analyses, except for LOESS, time from diagnosis was expressed as a discrete variable, with tests performed within several days after diagnosis imputed to the lowest number in the interval. Therefore, tests performed in the first 14 days since diagnosis are treated as “Day 0” and tests performed between day 360 and 449 (i.e. the last day observed, see **Table 1**) are treated as “Day 360”, see **Table S1**.

**Table 1.**
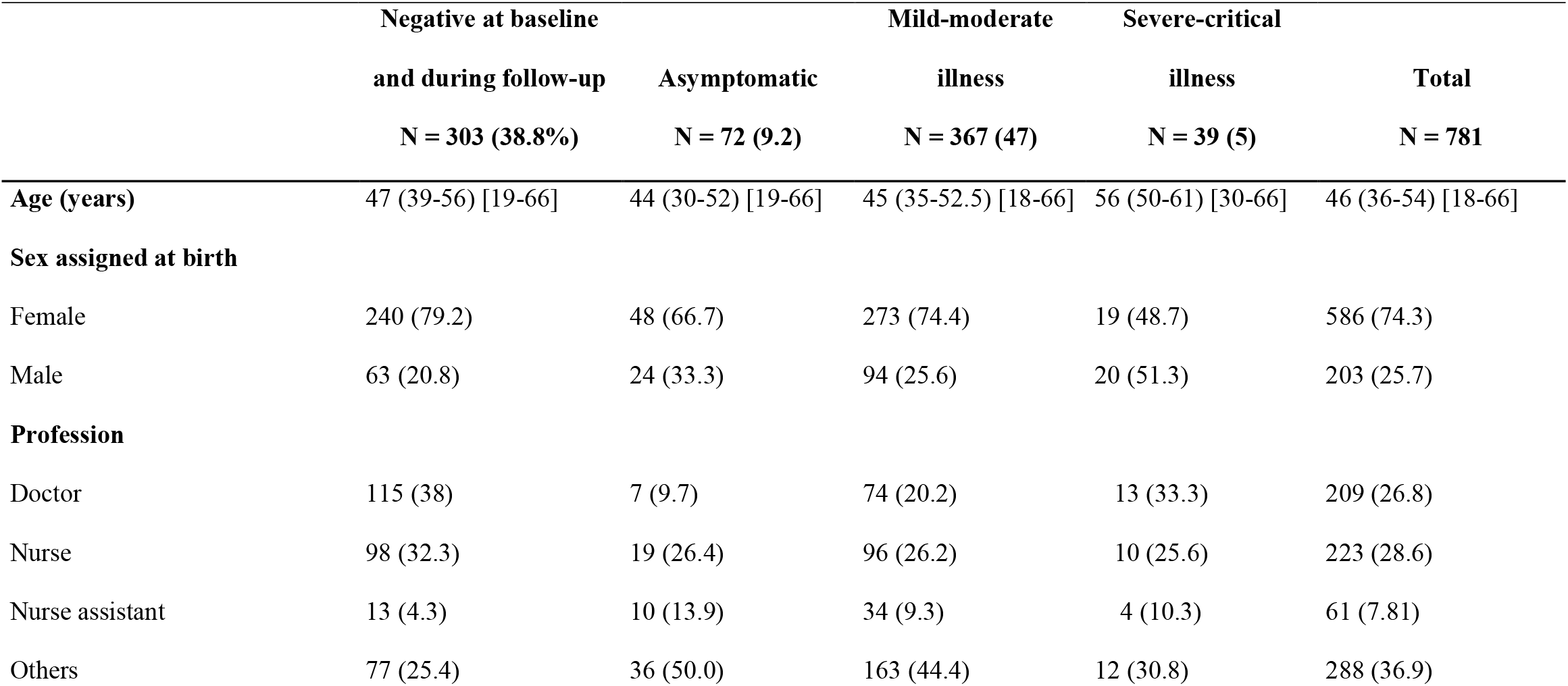
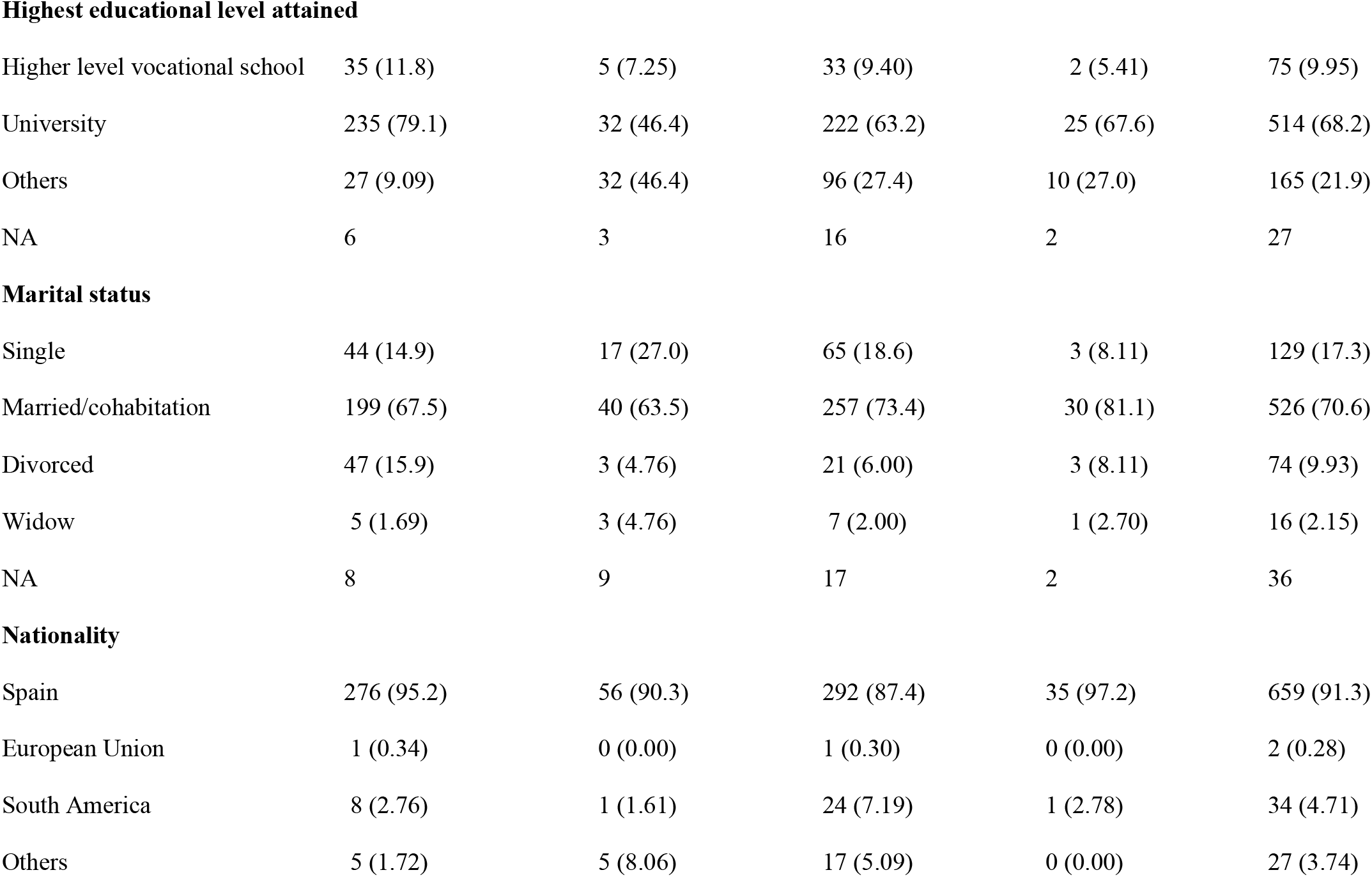
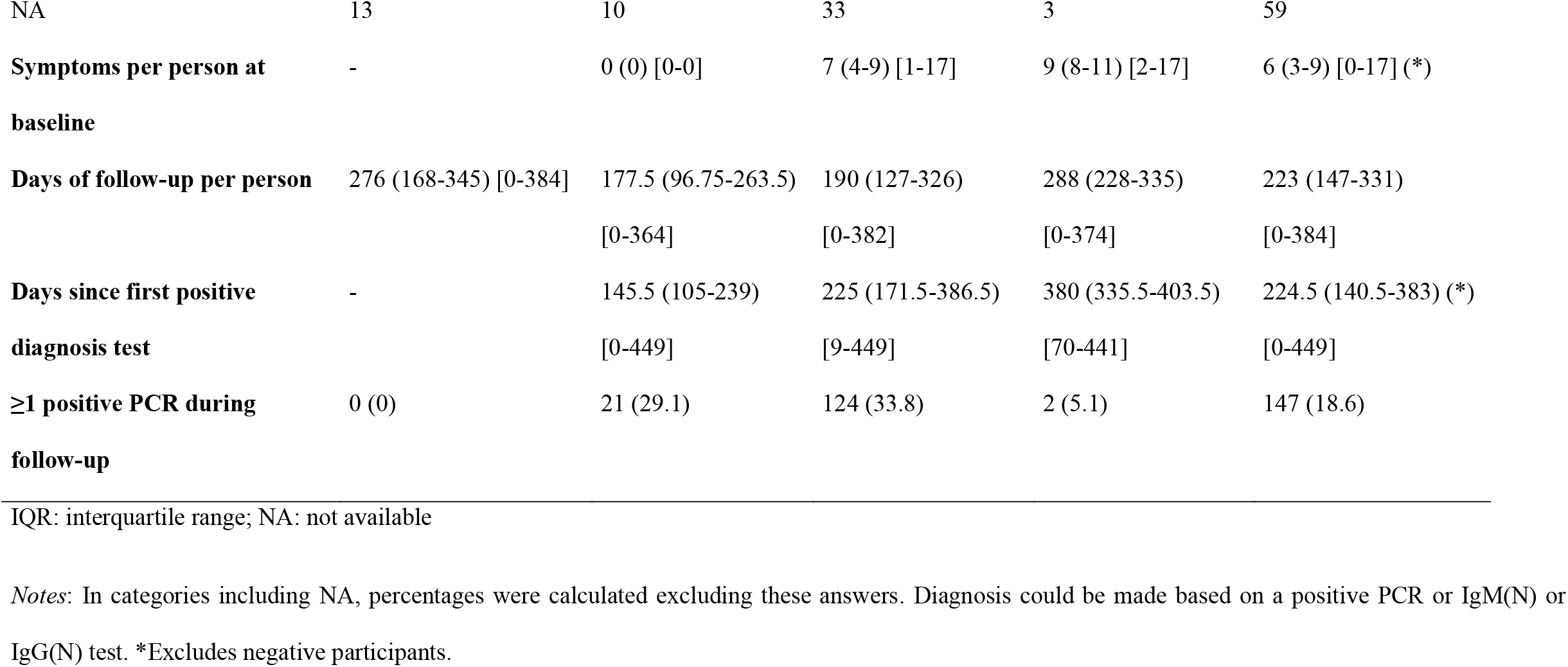
Demographic description and PCR testing for the study participants according to their clinical condition. Categoric variables are described as N (%), and numeric variables as median (IQR) [min, max].

|  | Negative at baseline<br>and during follow-up<br>N = 303 (38.8%) | Asymptomatic<br>N = 72 (9.2) | Mild-moderate<br>illness<br>N = 367 (47) | Severe-critical<br>illness<br>N = 39 (5) | Total<br>N = 781 |
| --- | --- | --- | --- | --- | --- |
| **Age (years)** | 47 (39-56) [19-66] | 44 (30-52) [19-66] | 45 (35-52.5) [18-66] | 56 (50-61) [30-66] | 46 (36-54) [18-66] |
| **Sex assigned at birth** |  |  |  |  |  |
| Female | 240 (79.2) | 48 (66.7) | 273 (74.4) | 19 (48.7) | 586 (74.3) |
| Male | 63 (20.8) | 24 (33.3) | 94 (25.6) | 20 (51.3) | 203 (25.7) |
| **Profession** |  |  |  |  |  |
| Doctor | 115 (38) | 7 (9.7) | 74 (20.2) | 13 (33.3) | 209 (26.8) |
| Nurse | 98 (32.3) | 19 (26.4) | 96 (26.2) | 10 (25.6) | 223 (28.6) |
| Nurse assistant | 13 (4.3) | 10 (13.9) | 34 (9.3) | 4 (10.3) | 61 (7.81) |
| Others | 77 (25.4) | 36 (50.0) | 163 (44.4) | 12 (30.8) | 288 (36.9) |
| **Highest educational level attained** |  |  |  |  |  |
| Higher level vocational school | 35 (11.8) | 5 (7.25) | 33 (9.40) | 2 (5.41) | 75 (9.95) |
| University | 235 (79.1) | 32 (46.4) | 222 (63.2) | 25 (67.6) | 514 (68.2) |
| Others | 27 (9.09) | 32 (46.4) | 96 (27.4) | 10 (27.0) | 165 (21.9) |
| NA | 6 | 3 | 16 | 2 | 27 |
| **Marital status** |  |  |  |  |  |
| Single | 44 (14.9) | 17 (27.0) | 65 (18.6) | 3 (8.11) | 129 (17.3) |
| Married/cohabitation | 199 (67.5) | 40 (63.5) | 257 (73.4) | 30 (81.1) | 526 (70.6) |
| Divorced | 47 (15.9) | 3 (4.76) | 21 (6.00) | 3 (8.11) | 74 (9.93) |
| Widow | 5 (1.69) | 3 (4.76) | 7 (2.00) | 1 (2.70) | 16 (2.15) |
| NA | 8 | 9 | 17 | 2 | 36 |
| **Nationality** |  |  |  |  |  |
| Spain | 276 (95.2) | 56 (90.3) | 292 (87.4) | 35 (97.2) | 659 (91.3) |
| European Union | 1 (0.34) | 0 (0.00) | 1 (0.30) | 0 (0.00) | 2 (0.28) |
| South America | 8 (2.76) | 1 (1.61) | 24 (7.19) | 1 (2.78) | 34 (4.71) |
| Others | 5 (1.72) | 5 (8.06) | 17 (5.09) | 0 (0.00) | 27 (3.74) |
| NA | 13 | 10 | 33 | 3 | 59 |
| **Symptoms per person at baseline** | - | 0 (0) [0-0] | 7 (4-9) [1-17] | 9 (8-11) [2-17] | 6 (3-9) [0-17] (*) |
| **Days of follow-up per person** | 276 (168-345) [0-384] | 177.5 (96.75-263.5) [0-364] | 190 (127-326) [0-382] | 288 (228-335) [0-374] | 223 (147-331) [0-384] |
| **Days since first positive diagnosis test** | - | 145.5 (105-239) [0-449] | 225 (171.5-386.5) [9-449] | 380 (335.5-403.5) [70-441] | 224.5 (140.5-383) (*) [0-449] |
| **≥1 positive PCR during follow-up** | 0 (0) | 21 (29.1) | 124 (33.8) | 2 (5.1) | 147 (18.6) |

IQR: interquartile range; NA: not available
*Notes:* In categories including NA, percentages were calculated excluding these answers. Diagnosis could be made based on a positive PCR or IgM(N) or IgG(N) test. \*Excludes negative participants.

All tests were two-sided. The significance level was set at p < 0.05. All analyses were performed with R version 4.0.0. See the Supplementary Appendix and Supplementary protocol.

## RESULTS

### Participant characteristics

A total of 860 participants were recruited, of whom 781 were eligible; 451 (57.7%) tested positive at baseline, and 27 (3.5%) had their first positive SARS-CoV-2 antibody test during follow-up (**Figure S1, Table 1**). Differences in the prevalence of specific clinical symptoms according to disease severity and sex are shown in **Table S2**.

### Evolution of seroprevalence in the ProHEpiC-19 study

**Table 2A** presents the participants with each possible combination of antibody test results, positive or negative, throughout follow-up. At baseline, more than one third of the participants (38.8%) tested negative for all antibodies, but this proportion decreased over time. From day 30 to day 180 of infection, more than 45% of participants tested positive for all antibodies. However, this proportion decreased from day 180 as the number of participants with positive IgM(N) values fell. By day 270, 12.1% of the participants were negative for all antibody tests, while 87.9% of the participants were at least IgG(N) or IgG(S) positive. As **Table 2B** shows, 67% of participants still had IgG(N) values over the positive threshold at day 360.

**Table 2.**
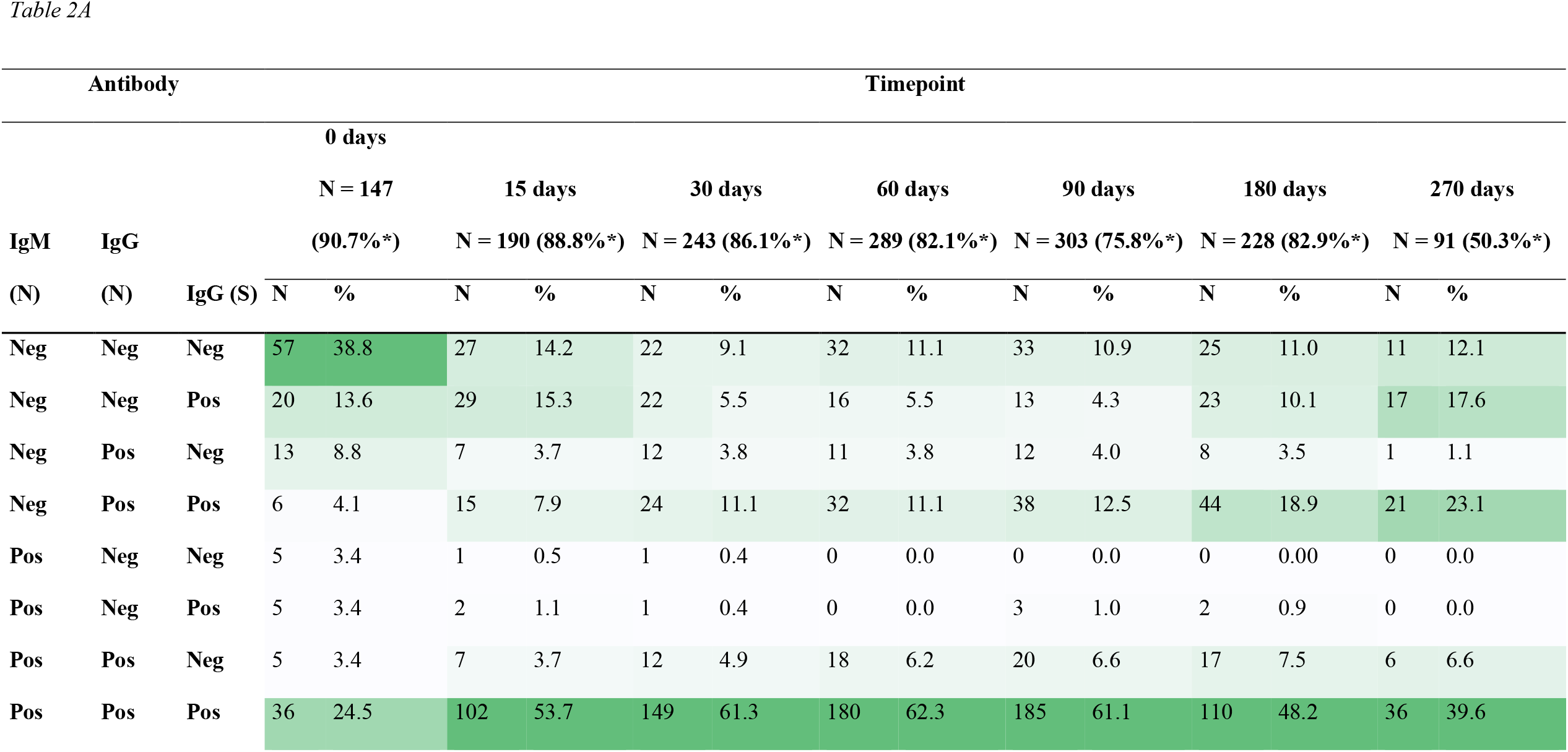
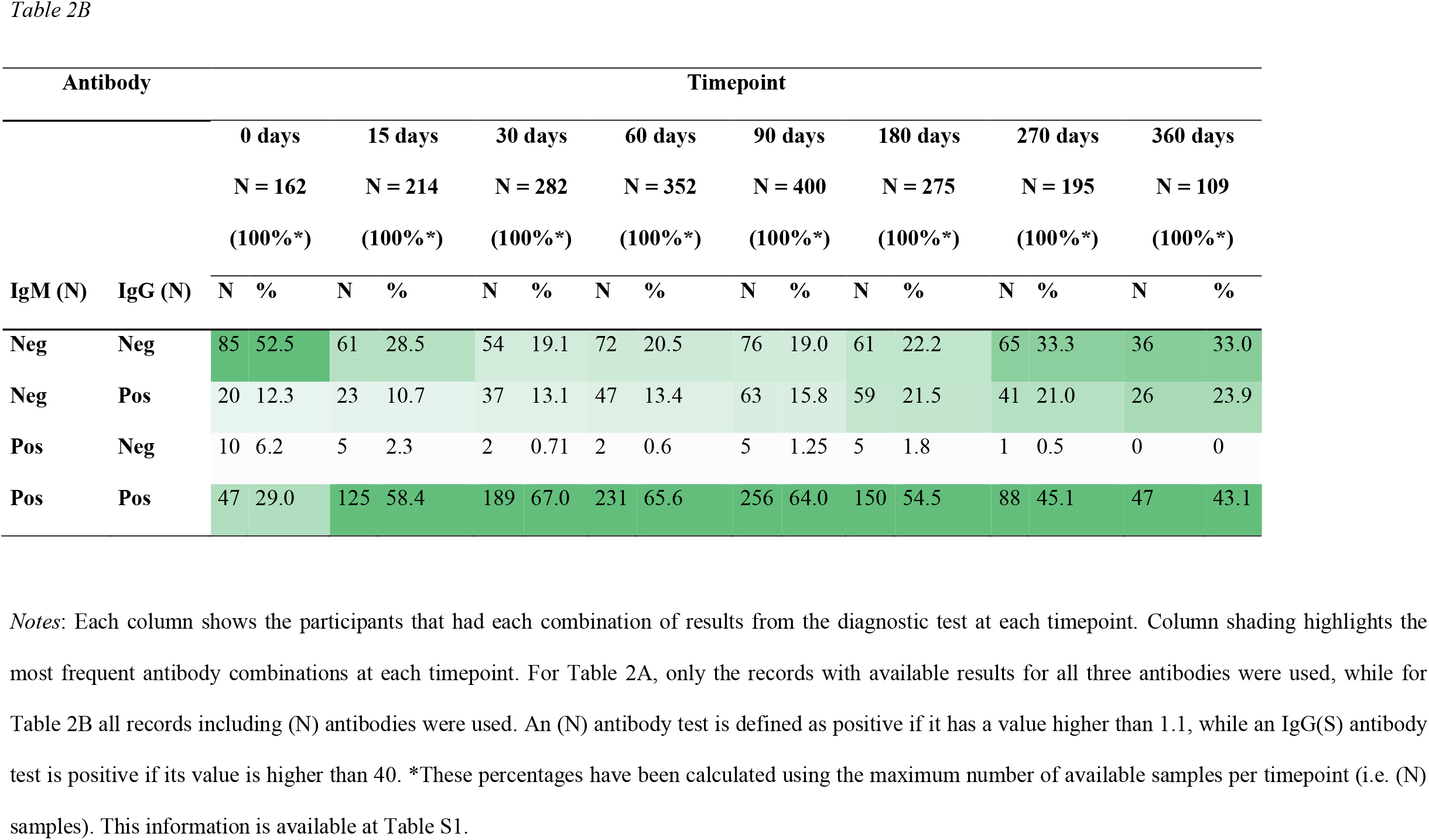
Description (N, %) of the results of the antibodies tests through the follow-up period.

### Levels of SARS-CoV-2 antibodies stratified by sex and disease severity

We found a statistically significant difference in all antibody levels between clinical conditions (P<0.001 both overall and pairwise comparisons). There was also a significant difference in antibody levels between males and females for all immunoglobulins: IgM(N) (P=0.015), IgG(N) (P<0.001), and IgG(S) (P=0.002).

Regarding the differences in antibody levels between the different time-points (**Figure 1**) median IgM(N) levels were below the threshold for positivity in people with asymptomatic and mild-moderate diseases after day 270 from diagnosis. However, IgG(N, S) levels still surpassed this threshold at day 360. The IgG(N) levels present a rise and fall from day 30, while IgG(S) levels remain practically constant following the first rise at day 15. Participants with severe disease consistently had higher levels of IgG (N, S) than patients with asymptomatic or mild-moderate disease. In terms of sex, there were no significant differences, except at day 30, and at day 60 only for IgG(S), when males showed higher levels than females.

**Figure 1.**
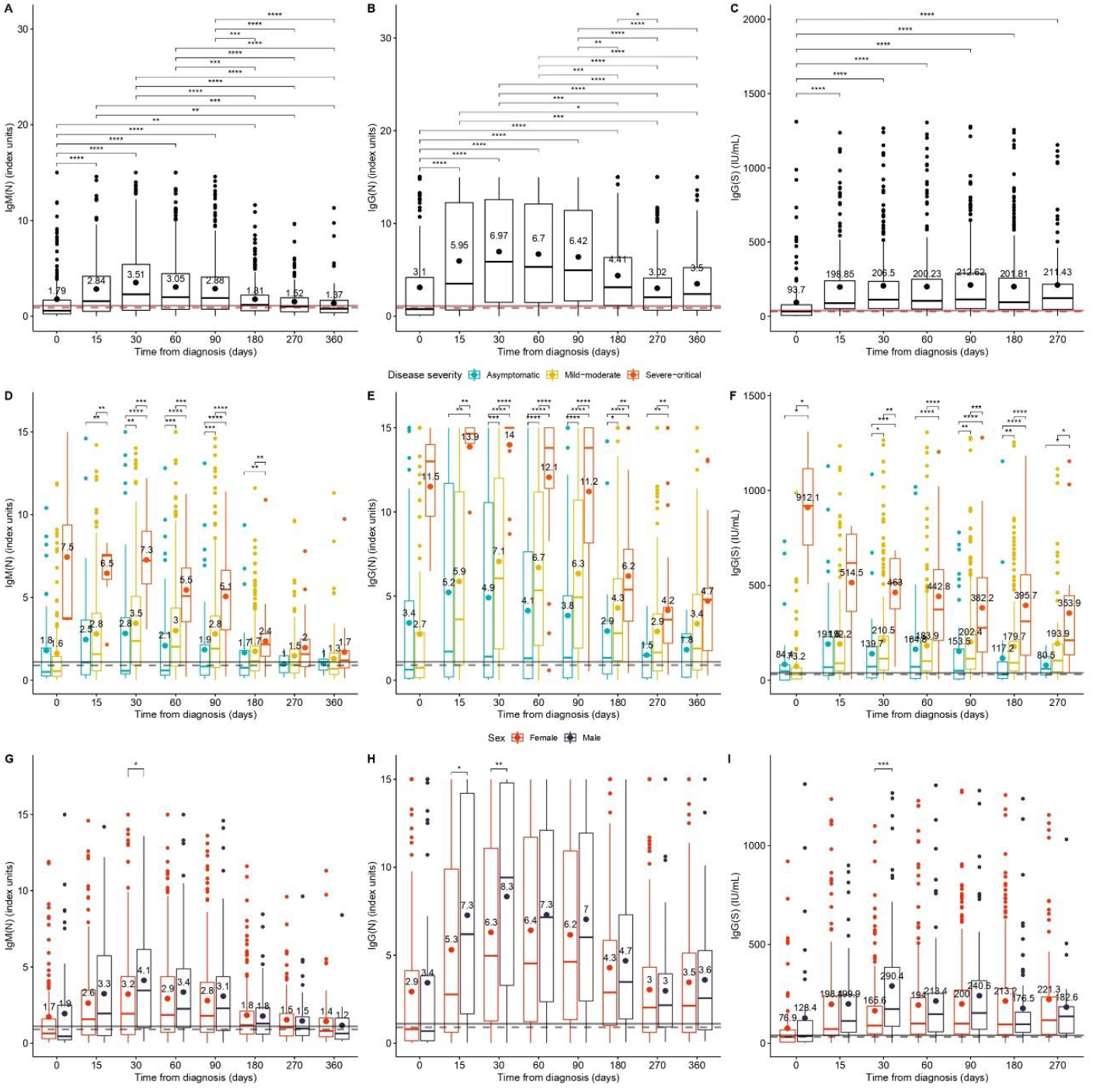
Description of the IgM (N), IgG (N) ang IgG(S) levels, by days since diagnosis. Antibody levels are represented with a boxplot together with a dot and text describing their mean value. The solid and dashed lines represent the limits of definition and uncertainty, respectively, for considering a positive result. Figures 1A-C show the significance of differences of the median antibody levels between days. Figures 1D-F show the significance of the median differences in antibody levels between clinical conditions within each day. Finally, Figures 1G-I show the significance of the median differences in antibody levels between sexes for each timepoint. A Holm-adjusted Dunn’s test was used in the first six figures, and a Holm-adjusted Mann-Whitney test in the last three. All the figures including (N) antibody results show a generalized rise in antibody levels up to day 30 since diagnosis, followed by a decline. Figures 1A and 1B show that both IgM(N) and IgG(N) levels from day 270 are no longer significantly different from those on day 0. Meanwhile, IgG(S) values (Figure 1C) remain constant after an initial rise on day 15. As for differences by clinical condition, throughout the follow-up period since diagnosis, the IgM(N), IgG(N) and IgG(S) levels (see Figure 1D-F, respectively) of patients with severe-critical disease were higher almost to the end of follow-up. Although the IgM(N) and IgG(N) levels of asymptomatic patients were initially higher than those of mild-moderate patients, from day 15 they started to decline. From day 270 onwards, the median IgM(N) values of asymptomatic and mild to moderate patients were already below the threshold considered positive. In the case of median IgG(N) levels, although their decrease was evident from day 60 onwards, on day 360 since diagnosis they were still above the positive threshold across the clinical spectrum. On the other hand, median levels of IgG(S) remained virtually constant throughout the entire follow-up period, regardless of disease severity. Compared to women, men had higher levels of all antibodies analyzed on day 30 and of IgG(N) on day 60.

### Dynamics of IgM and IgG reveal differential kinetics

The kinetics for the IgG isotopes varied between N and S. In relation to the kinetics of the three antibodies stratified by disease severity, both LOESS (**Figures 2A, 2C, 2D**) and NLME (**Figures 2B, 2D, 2E**) adjustment methods showed a general rise in both IgM(N) and IgG(N) levels up to day 30, followed by a decay whose rate depended on disease severity. IgG(S) levels increased at day 15 and remained relatively constant over time.

**Figure 2.**
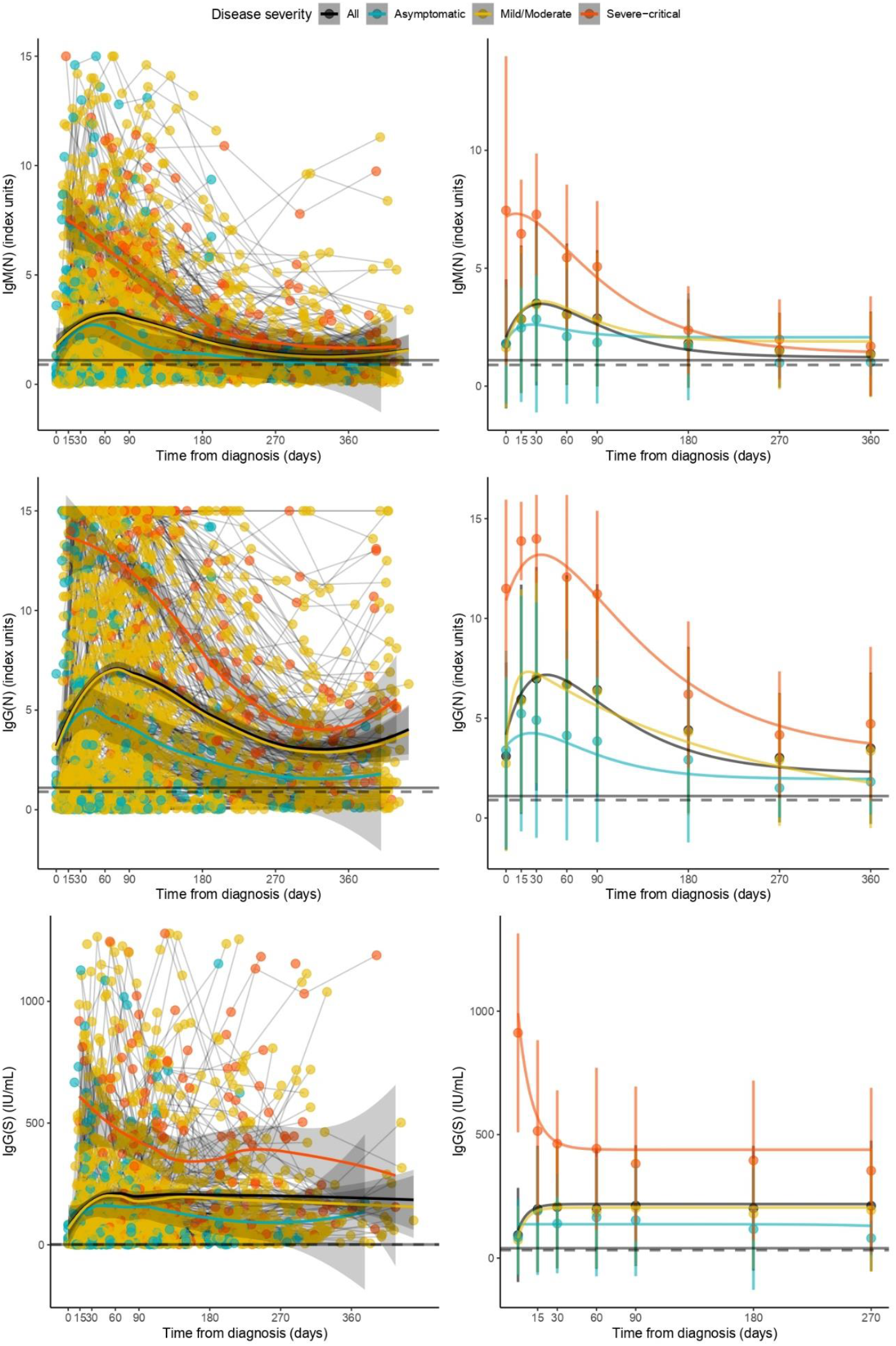
Decay of IgM(N), IgG(N) and IgG(S) levels since diagnosis in total sample and by clinical condition. Figures 2A, 2C and 2E show LOESS regression models with all available data, connecting results belonging to the same participants. Figures 2B, 2D and 2F show the estimated non-linear mixed-effect (NLME) model curves. Each point corresponds to the mean value at each timepoint. The bars show the mean ± standard deviation interval (NLME). The solid and dashed lines represent the limits of definition and uncertainty, respectively, for considering a positive result. Overall, the level of the antigen response correlated with the severity of the clinical presentation.

**Table S3** shows the parameters estimated for each component of the NLME curves. For both IgM(N) and IgG(N), levels were significantly higher in severe-critical participants at the beginning of the study, the increase rate was slower in asymptomatic disease, and the decrease rate in both asymptomatic and severe-critical disease was faster than in mild-moderate cases. Regarding IgG (S) levels, severe-critical participants showed significantly higher values on day 270 than mild-moderate participants. Asymptomatic participants had significantly lower values on day 270, but they were still positive. However, as can be seen in **Figure 2F**, the levels were practically constant in the three groups.

**Figure 3** presents the kinetics stratified by sex. The evolution patterns of the three antibodies are practically identical except for a peak in men on day 30. Then, the increase rate of IgM(N) levels is higher in males. The parameters of these NLME curves can be also found in **Table S3**.

**Figure 3.**
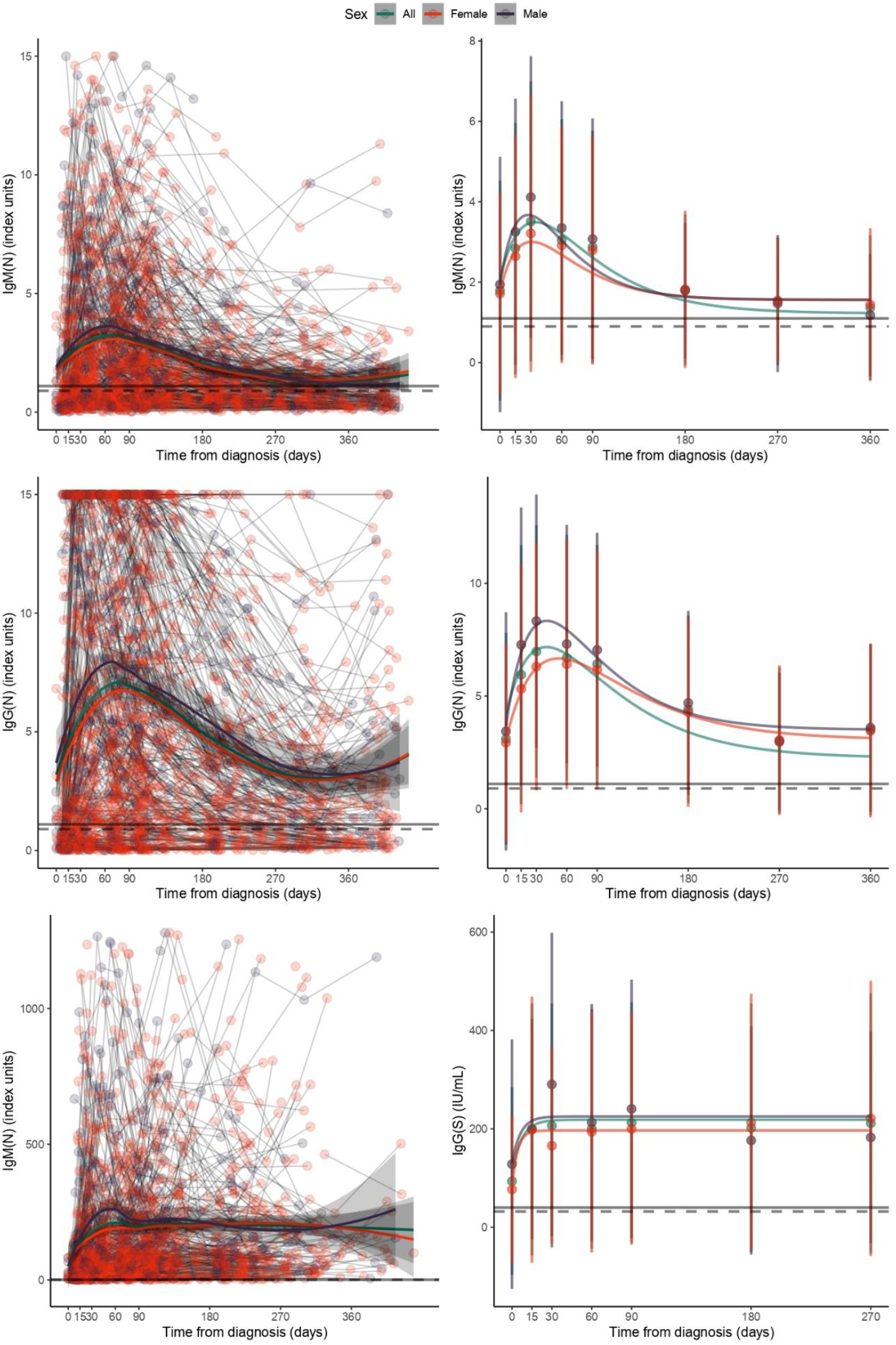
Decay of IgM(N), IgG(N) and IgG(S) levels since diagnosis, both aggregated and stratified by sex. Figures 3A, 3C and 3E show LOESS regression models with all available data, connecting results belonging to the same participants, as well as the LOESS curves. Figures 3B, 3D and 3F show the estimated non-linear mixed-effect (NLME) model curves. Each point corresponds to the mean value at each timepoint. The bars show the mean ± standard deviation interval (NLME). The solid and dashed lines represent the limits of definition and uncertainty, respectively, for considering a positive result. Overall, the evolution of antibody levels was very similar between sexes, with the only difference being a peak in antibody levels in males in day 30 from diagnosis.

## DISCUSSION

Serological response to SARS-CoV-2 differs along the clinical spectrum. Our results show that antibody response starts within 15 days of infection for the three studied isotypes. Thereafter, their behavior diverges according to disease severity. We found a higher level of antibodies in patients with severe versus asymptomatic infections. Likewise, our results corroborate the early appearance of IgG(N) and (S) ^14, 6^.

Asymptomatic individuals maintain antibody levels above the threshold for IgM(N) up to six months, while antibodies for IgG(N) and (S) remain above the threshold to day 360 and 270, respectively. The long-term presence of IgG(N) antibodies and their efficacy needs further research ^15, 16^. Compared with asymptomatic cases, participants with mild-moderate COVID-19 presented higher IgG antibody levels for the entire follow-up, and their IgM (N) levels remained above the threshold at least until 360 days post-infection. These findings are similar to those reported at six months in a longitudinal study of IgM(S) and can be explained by the differentiation of B cells to IgM memory plasma cells that continue to produce IgM isotype antibodies for at least a year ^17^. Another recent study also found positive IgM levels for up to one year, although the isotopes were different: IgM (S, receptor binding domain). Severe-critical participants had higher antibody levels than the other groups in the first 30 days of follow-up and maintained the highest levels for all three isotopes throughout the entire follow-up period. This is consistent with the results of other studies conducted over a shorter period of time ^18, 9^.

The LOESS models enable a description of the trajectories of the antibody levels, while the NLME facilitates the comparison of the average trajectories according to disease severity, accounting for the non-linearity of the antibody levels. The model’s goodness-of-fit confirms its validity and explanatory capacity, making ours the first study allowing an estimate of antibody values against SARS-CoV-2, with potential applications in vaccinated people to adjust (re)vaccination criteria. Furthermore, the kinetic model of IgG (S) can be easily adapted for estimating antibody levels using other units by applying the corresponding conversion factors. Likewise, these models can be adapted to the kinetics calculated in vaccinated people, informing the criteria for revaccination.

This study describes, for the first time to our knowledge, differential, and specific dynamics of IgG(N) and IgG(S) production after SARS-CoV-2 infection with high resolution. IgG(N) shows a rapid rise and a differential downward slope depending on disease severity, stabilizing between day 270 and 360. In contrast, IgG(S) kinetics shows a flat response from day 30 post-infection. Differences have been observed elsewhere ^19^ and may be attributable to the differences between the S and N proteins of SARS-COV-2 in the molecular structure, amounts in the viral particle, and specific functions. While the former facilitates the entry of the virus into the host cell, the latter has a role in viral genomic packaging ^20^. Monitoring of both IgG N and S antibodies can help identify stimulation of memory plasma cells by two different antigens. The maintenance of an N and S polyclonal IgG response may protect against possible reinfections and immunological escape from the vaccination. Further in-depth immunological studies on IgG N and S specificities will be important to address their role in protection from severity and reinfection in natural infection and vaccination settings.

Regarding sex, humoral immune response behavior is the same in men and women, although IgG(N) or IgG(S) levels or titers are always higher in men. Other studies at eight months’ follow-up indicate that sex and severe disease are associated with differences in immune memory to SARS-CoV-2. However, most of the heterogeneity in immune memory to SARS-CoV-2 is still unexplained, and further investigation on the role of cellular immunity and memory responses is needed in particular groups, (non-seroconvertors, people with immunodeficiencies, and autoimmune disorders) ^2, 21^.

Anti-SARS CoV2 antibodies were determined by ELISA techniques, although anti-N antibodies were semi-quantitative and anti-S antibodies, quantitative (IgG). The maximum levels of IgM(N) and IgG(N) antibodies tested could be higher, as the technique used was semi-quantitative, and the index calculation reached a maximum value of 15. Semi-quantitative tests have some limitations, especially at the upper threshold, where further dilutions may be necessary. The limited availability of quantitative in vitro diagnostic techniques at the beginning of the pandemic necessitated the use of semi-quantitative techniques. Also, given the rapid development of diagnostic tests and the lack of information on them, we had to conduct an ELISA evaluation study prior to this work. In May 2020, when the study began, the techniques for measuring antibodies to SARS-CoV-2 were qualitative or semi-quantitative, so once the WHO international standard for quantification of IgG(S) antibodies was established we retrospectively analyzed a measurement of IgG spike S1 using a quantitative technique. The follow-up of IgG(S) levels stopped as participants were vaccinated, while the IgM(N) and IgG(N) continued.

Key questions remain unanswered, such as whether these models kinetics will be valid in vaccinated individuals; if the kinetics and duration of anti-S antibodies are similar in natural infection and vaccination; and whether previously infected and uninfected patients will show the same kinetics after vaccination. Epidemiological modelling studies, including long-term immune monitoring, will be crucial in the case of SARS CoV-2 but also to evaluate the interactions with other coronaviruses for accurate predictions in the case of other viral coinfections (e.g. flu, other coronavirus, HIV-1) ^22, 23^

To conclude, we monitored three antibodies for just over a year, analyzing their titers and kinetics according to clinical severity and sex. NLME models helped explain the average trajectory across the clinical spectrum at one year, confirming that infected people maintain immunity with IgG isotypes. Larger studies with a longer follow-up period are still necessary. The kinetic models defined in this study with quantitative IgG(S) determinations can serve as a reference point to indicate when infected people should be (re)vaccinated and monitor the vaccinated population.

## Data Availability

The study data will not be openly available. If researchers or institutions are interested in the data, the authors will be able to release the data after asking the study participants for their consent and signing the relevant legal agreements allowing the data to be released to third parties.

## Acknowledgment

The authors would like to sincerely thank the participants for their effort and selfless involvement in the ProHEpiC-19 cohort study. Also, they want to thank the Management Department, Primary Care Directorate and the Directorate of the Clinical Laboratory of the Metropolitan North for the facilities they have given for the development of the project. In addition, we deeply thank the technical staff of IrsiCaixa for the processing of samples (L. Ruiz, E. Grau, R. Ayen, L. Gomez, C. Ramirez, M. Martinez, T Puig. R Penya, O Blanch-Lombarte).

ProHEPiC-19 was supported by the Health Ministry of Generalitat de Catalunya, Call COVID19-PoC number SLT16_04 and supported in part by grants from National Health Institute Carlos III (ISCIII) COV20/00660, and RETIC RD16/0025/0041 (Co-funded by European Regional Development Fund/European Social Fund) for Julia Garcia Prieto. The funders had no role in study design, data collection and analysis, the decision to publish or drafting of the manuscript. We thank “CERCA Programme/Generalitat de Catalunya” for institutional support.

---

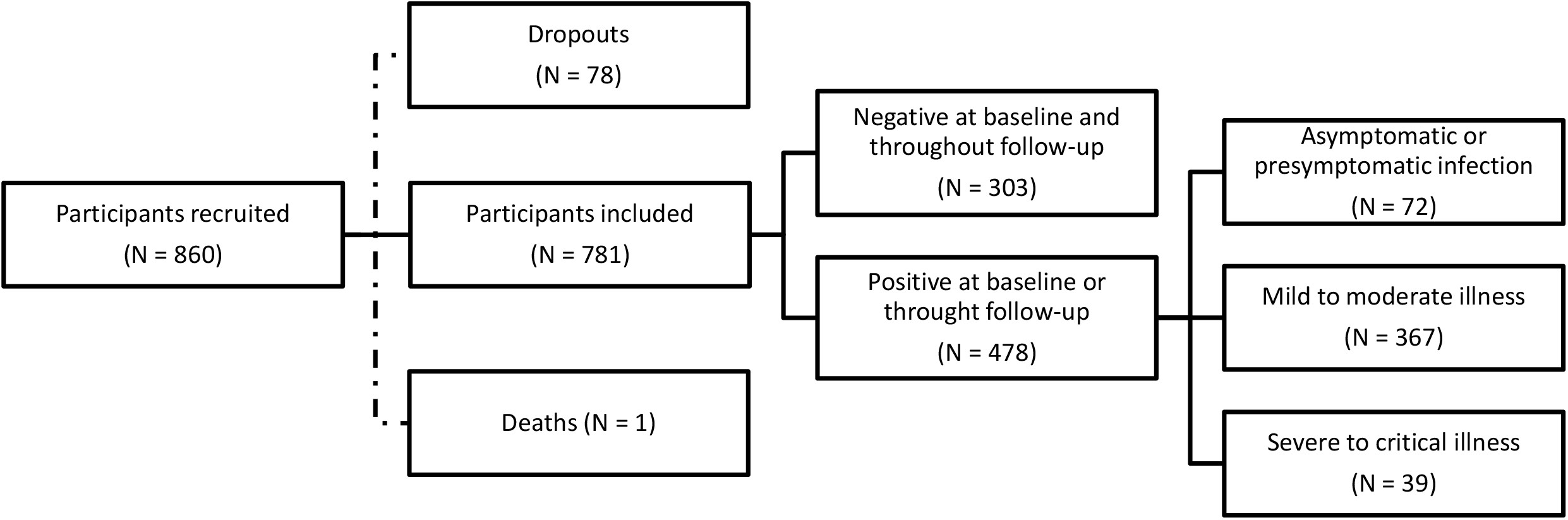

## REFERENCES

1. Galit and Seder RGA. The Power of Antibody-Based Surveillance. N Engl J Med 2020;383(18):1782–4.

2. Dan JM, Mateus J, Kato Y, et al. Immunological memory to SARS-CoV-2 assessed for up to 8 months after infection. Science (80-) 2021;371(6529).

3. Trinité B, Tarrés-Freixas F, Rodon J, et al. SARS-CoV-2 infection elicits a rapid neutralizing antibody response that correlates with disease severity. Sci Rep 2021;11(1).

4. Carrillo J, Izquierdo-Useros N, Ávila-Nieto C, Pradenas E, Clotet B, Blanco J. Humoral immune responses and neutralizing antibodies against SARS-CoV-2; implications in pathogenesis and protective immunity. Biochem Biophys Res Commun 2021;538:187–91.

5. Wang P, Liu L, Nair MS, et al. SARS-CoV-2 neutralizing antibody responses are more robust in patients with severe disease. Emerg. Microbes Infect. 2020;9(1):2091–3.

6. Long QX, Tang XJ, Shi QL, et al. Clinical and immunological assessment of asymptomatic SARS-CoV-2 infections. Nat Med 2020;26(8):1200–4.

7. Gudbjartsson DF, Norddahl GL, Melsted P, et al. Humoral Immune Response to SARS-CoV-2 in Iceland. N Engl J Med 2020;383(18):1724–34.

8. Lumley SF, O’Donnell D, Stoesser NE, et al. Antibody Status and Incidence of SARS-CoV-2 Infection in Health Care Workers. N Engl J Med 2021;384(6):533–40.

9. Huang AT, Garcia-Carreras B, Hitchings MDT, et al. A systematic review of antibody mediated immunity to coronaviruses: kinetics, correlates of protection, and association with severity. Nat Commun 2020;11(1).

10. Dobaño C, Ramírez-Morros A, Alonso S, et al. Persistence and baseline determinants of seropositivity and reinfection rates in health care workers up to 12.5 months after COVID-19. BMC Med [Internet] 2021;19(1):155. Available from: https://bmcmedicine.biomedcentral.com/articles/10.1186/s12916-021-02032-2

11. Kirtipal N, Bharadwaj S, Kang SG. From SARS to SARS-CoV-2, insights on structure, pathogenicity and immunity aspects of pandemic human coronaviruses. Infect Genet Evol 2020;85:104502.

12. Walsh EE, Frenck RW, Falsey AR, et al. Safety and Immunogenicity of Two RNA-Based Covid-19 Vaccine Candidates. N Engl J Med 2020;383(25):2439–50.

13. Mattiuzzo G, Bentley EM, Hassall M, Routley S. Establishment of the WHO International Standard and Reference Panel for anti-SARS-CoV-2 antibody [Internet]. 2020. Available from: https://www.who.int/publications/m/item/WHO-BS-2020.2403

14. Fu Y, Pan Y, Li Z, Li Y. The Utility of Specific Antibodies Against SARS-CoV-2 in Laboratory Diagnosis. Front. Microbiol. 2021;11.

15. Harrington WE, Trakhimets O, Andrade D V., et al. Rapid decline of neutralizing antibodies is associated with decay of IgM in adults recovered from mild COVID-19. Cell Reports Med 2021;2(4).

16. Zhao J, Zhao S, Ou J, et al. COVID-19: Coronavirus Vaccine Development Updates. Front Immunol 2020;11(December):1–19.

17. Turner JS, Kim W, Kalaidina E, et al. SARS-CoV-2 infection induces long-lived bone marrow plasma cells in humans. Nature 2021;

18. Graham NR, Whitaker AN, Strother CA, et al. Kinetics and isotype assessment of antibodies targeting the spike protein receptor-binding domain of severe acute respiratory syndrome-coronavirus-2 in COVID-19 patients as a function of age, biological sex and disease severity. Clin Transl Immunol 2020;9(10).

19. Alfego D, Sullivan A, Poirier B, Williams J, Adcock D, Letovsky S. A population-based analysis of the longevity of SARS-CoV-2 antibody seropositivity in the United States. EClinicalMedicine 2021;36:100902.

20. Mariano G, Farthing RJ, Lale-Farjat SLM, Bergeron JRC. Structural Characterization of SARS-CoV-2: Where We Are, and Where We Need to Be. Front. Mol. Biosci. 2020;7.

21. Kilpeläinen A, Jimenez-Moyano E, Blanch-Lombarte O, et al. Highly functional Cellular Immunity in SARS-CoV-2 Non-Seroconvertors is associated with immune protection 5. bioRxiv [Internet] 2021;2021.05.04.438781. Available from: https://doi.org/10.1101/2021.05.04.438781

22. Petersen E, Koopmans M, Go U, et al. Comparing SARS-CoV-2 with SARS-CoV and influenza pandemics. Lancet Infect Dis 2020;20:e238–44.

23. Fischer W Giorgi, Elena E Chakraborty, Srirupa Nguyen, Kien Bhattacharya, Tanmoy Theiler J, Goloboff PA, et al. HIV-1 and SARS-CoV-2: Patterns in the evolution of two pandemic pathogens. Cell Host Microbe 2021;29:1093–100.

