## Supplementary Material for "Antibody kinetics to SARS-CoV-2 at 13.5 months, by disease severity"

### Supplementary Methods

#### 1 INDEX

#### 2 PARTICIPANT INCLUSION

Participants recruited prior to testing positive who seroconverted during the follow-up period (N = 27 (3.4%)) were only considered in the analysis of positive participants from the time of diagnosis. The temporal relationship between the time of diagnosis and the first study visit of the 478 SARS-CoV-2 positive participants is presented in the table below. The number of days from diagnosis was discretized as explained in Section 2.2. of this appendix.

| **Timepoint (days from diagnosis)** | **Patients joining the trial (N)** | **Frequency** | **Cumulative frequency** |
|---|---|---|---|
| **0** | 162 | 33.9 | 33.9 |
| **15** | 82 | 17.2 | 51 |
| **30** | 63 | 13.2 | 64.2 |
| **60** | 102 | 21.3 | 85.6 |
| **90** | 47 | 9.8 | 95.4 |

| | | | |
|---|---|---|---|
| **180** | 19 | 4 | 99.4 |
| **270** | 3 | 0.6 | 100 |
| **360** | 0 | 0 | 100 |

### 3 Statistical analysis

An outlier detection was performed on IgG(S) antibody levels, considering outliers the values belonging to the most extreme 0.3%, according to the z-score. Sixty-five IgG(S) values were thus excluded. No outlier detection was performed on (N) antibodies as their value was limited ([0, 15]).

All tests were two-sided. Significance level was set to $p < 0.05$. All analyses were performed using R version 4.0.4 or higher. The code that produced all the results can be found at https://github.com/IDIAPJGol/ProHEpic_Antibodies.

#### 3.1 Statistical tests

We used the chi-squared test to identify significant differences between the expected and the observed frequencies for categorical comparisons (e.g., the prevalence of symptoms according to biological sex or disease severity).

To evaluate differences in SARS-CoV-2 antibody (IgM, IgG) levels across the clinical spectrum, we used a Kruskal-Wallis test after assessing the normality of the distribution (Shapiro-test p-value < 0.05). This test studies the differences in the medians between the different categories, using all available antibody results. A post-hoc analysis was performed using a Holm-adjusted Dunn's test to find significant differences. Similarly, a Holm-adjusted Mann-Whitney test was performed to look for differences in antibody levels between sexes.

#### 3.2 Descriptive analysis of the evolution of antibody levels from diagnosis

An initial descriptive analysis of the evolution of antibodies levels since diagnosis was performed by means of boxplots and statistical tests. Time from diagnosis was discretized as follows to group test results with a temporal relationship: tests done in the first 14 days from diagnosis were treated as "day 0"; tests from days 15 to 29, as "day 15"; from days 30 to 59, as "day 30"; from days 60 to 89, as "day 60"; from days 90 to 179, as "day 90"; from days 180 to 269, as "day 180"; from days 270 to 360, as "day 270"; and from days 360 up to end of follow-up, as “day 360”. The number of available samples for each timepoint and antibody is presented in Table S2.

Due to the very low proportion of participants arriving at day 360 of follow-up without having been vaccinated and the outlier removal, the number of available samples of IgG(S) antibody levels was very low, and they were excluded from further analysis.

Once the time was discretized, boxplots showing the distribution of the IgM and IgG (both N and S) values per day were obtained. We obtained both an overall distribution for each timepoint and a separate distribution by disease severity and sex. We used the former to compare the distributions of the timepoints with each other, seeking to find significant differences using a Holm-adjusted Dunn's test, and the latter to compare distributions within each timepoint to find differences between groups, either by sex or disease severity. For the comparison between disease severity each timepoint, a Holm-adjusted Dunn’s test was used, while for the comparison between sex, we used a Holm-adjusted Mann-Whitney test.

Locally estimated scatterplot smoothing (LOESS) models were fit as a second descriptive approach to study the evolution of both IgM and IgG antibodies from diagnosis day. LOESS regression consists of fitting simple models (e.g., first- or second-degree polynomials) in subsets of data determined by a nearest-neighbors algorithm. Figures 3 and S1 show the LOESS curves, stratified either by disease severity or sex, together with their 95% confidence interval (CI). We used all the data available except for participants who never tested positive, grouping the data points per participant in the figures.

##### 3.3 Nonlinear mixed-effects (NLME) models

Mixed-effects models are generally useful when there are multiple measures per participant, as in this study. Random-effects are included in the model to test if they may be causing any significant, and generally unexpected, influence in the modeling built by the fixed effects. Therefore, introducing random effects in the model helps to control unobserved variance.

In this study, we used NLME models to study possible differences in the dynamics (kinetics) of SARS-CoV-2 antibodies (IgM, IgG) levels from the time of diagnosis day. After fitting a general model, the differences were studied between disease severity and sex. "Mild to moderate illness" and "Female" were used as reference groups, respectively, as they were the largest groups. To fit these models, we used all the data available except for those from participants who never tested positive. Time from diagnosis was discretized as for the first descriptive analysis.

The general equations to be fitted to model mean values of antibodies at a time ***t*** was:

$$IgM(N)\ (t) = b_1 + (b_0 - b_1)e^{-k_1 t} - b_1 k_1 (e^{-k_1 t} - e^{-k_2 t})$$

$$IgG(N)\ (t) = b_1 + (b_0 - b_1)e^{-k_1 t} - b_1 k_1 (e^{-k_1 t} - e^{-k_2 t})$$

$$IgG(S)\ (t) = b_1 + (b_0 - b_1)e^{-k_1 t} - b_1 k_1 e^{-k_1 t}$$

Normalizing $t$ according to the maximum length of follow-up (i.e., 360 for (N) antibodies, 270 for (S) antibody). This general equation corresponds to an equation describing an exponential rise followed by an exponential decay, where $b_0$ corresponds to the baseline value, $b_1$ to the asymptotic value (i.e., antibody levels at $t \rightarrow \infty$), $k_1$ to the rise rate, and $k_2$ to the decay rate.

Nonlinear mixed-effects models were built using the nlme package (Pinheiro J. et al., 2020). This package needs to have prior values for the parameters. The following table shows the prior values used to start the models. The final values of the parameters after fitting the model can be seen in Table S3.

| SARS-CoV-2 antibody | Model | Parameter | Prior value |
|---|---|---|---|
| IgM (N) | Complete | b0 - Baseline value | 1.8 |
| | | b1 - Asymptotic value (i.e., antibody levels at t →∞) | 1.3 |
| | | $k_1$ - Rise rate | 6 |
| | | $k_2$ - Decay rate | 12 |
| | IgM (N) ~ Disease severity | $b_0$ (Mild-moderate) | 1.6 |
| | | $b_0$ (Asymptomatic) | 0 |
| | | $b_0$ (Severe-critical) | 0 |
| | | $b_1$ (Mild-moderate) | 1.3 |
| | | $b_1$ (Asymptomatic) | 0 |
| | | $b_1$ (Severe-critical) | 0 |
| | | $k_1$ (Mild-moderate) | 10 |
| | | $k_1$ (Asymptomatic) | 0 |
| | | $k_1$ (Severe/critical) | 0 |
| | | $k_2$ (Mild-moderate) | 6 |
| | | $k_2$ (Asymptomatic) | 0 |
| | | $k_2$ (Severe-critical) | 0 |
| | IgM (N) ~ Sex | $b_0$ (Female) | 1.7 |
| | | $b_0$ (Male) | 0 |
| | | $b_1$ (Female) | 1.4 |
| | | $b_1$ (Male) | 0 |
| | | $k_1$ (Female) | 12 |
| | | $k_1$ (Male) | 0 |

| | | | |
|---|---|---|---|
| | | $k_2$ (Female) | 6 |
| | | $k_2$ (Male) | 0 |
| IgG (N) | **Complete** | b0 - Baseline value | 3 |
| | | b1 - Asymptotic value (i.e., antibody levels at t →∞) | 3.5 |
| | | $k_1$ - Rise rate | 6 |
| | | $k_2$ - Decay rate | 12 |
| | **IgG (N) ~ Disease severity** | $b_0$ (Mild-moderate) | 3 |
| | | $b_0$ (Asymptomatic) | 0 |
| | | $b_0$ (Severe-critical) | 0 |
| | | $b_1$ (Mild-moderate) | 3.5 |
| | | $b_1$ (Asymptomatic) | 0 |
| | | $b_1$ (Severe-critical) | 0 |
| | | $k_1$ (Mild-moderate) | 6 |
| | | $k_1$ (Asymptomatic) | 0 |
| | | $k_1$ (Severe-critical) | 0 |
| | | $k_2$ (Mild-moderate) | 12 |
| | | $k_2$ (Asymptomatic) | 0 |
| | | $k_2$ (Severe-critical) | 0 |
| | **IgG (N) ~ Sex** | $b_0$ (Female) | 3 |
| | | $b_0$ (Male) | 0 |
| | | $b_1$ (Female) | 3.5 |
| | | $b_1$ (Male) | 0 |
| | | $k_1$ (Female) | 6 |
| | | $k_1$ (Male) | 3 |

| | | | |
|---|---|---|---|
| | | $k_2$ (Female) | 12 |
| | | $k_2$ (Male) | 2 |
| **IgG (S)** | **Complete** | b0 - Baseline value | 90 |
| | | b1 - Asymptotic value (i.e., antibody levels at t →∞) | 210 |
| | | $k_1$ - Rise rate | 26 |
| | **IgG (S) ~ Disease severity** | $b_0$ (Mild-moderate) | 70 |
| | | $b_0$ (Asymptomatic) | 10 |
| | | $b_0$ (Severe-critical) | 900 |
| | | $b_1$ (Mild-moderate) | 200 |
| | | $b_1$ (Asymptomatic) | 0 |
| | | $b_1$ (Severe-critical) | 150 |
| | | $k_1$ (Mild-moderate) | 30 |
| | | $k_1$ (Asymptomatic) | 0 |
| | | $k_1$ (Severe-critical) | 0 |
| | **IgG (S) ~ Sex** | $b_0$ (Female) | 80 |
| | | $b_0$ (Male) | 40 |
| | | $b_1$ (Female) | 240 |
| | | $b_1$ (Male) | -40 |
| | | $k_1$ (Female) | 24 |
| | | $k_1$ (Male) | 0 |

The adjusted **general** equations are:

$$IgM(N)\ (t) = 1.223 + 0.848e^{-10.754t} - 13.152(e^{-10.754t} - e^{-7.103t})$$

$$IgG(N)\ (t) = 2.243 + 1.924e^{-9.723t} - 21.809(e^{-9.723t} - e^{-5.619t})$$

$$IgG(S)\ (t) = 218.623 + 7983.968e^{-37.008t} - 8106.54e^{-37.008t}$$

Normalizing *t* according to the maximum length of follow-up (i.e., 360 for (N) antibodies, 270 for (S) antibody).

The adjusted equations for patients having **asymptomatic disease** are:

$$IgM(N)\ (t) = 2.075 - 0.146e^{-15.372t} - 31.897(e^{-15.372t} - e^{-14.621t})$$

$$IgG(N)\ (t) = 1.951 + 1.641e^{-9.866t} - 19.249(e^{-9.866t} - e^{-7.827t})$$

$$IgG(S)\ (t) = 137.139 - 1163.11e^{8.481t} + 1163.08e^{8.481t}$$

Normalizing *t* according to the maximum length of follow-up (i.e., 360 for (N) antibodies, 270 for (S) antibody).

The adjusted equations for patients with **mild to moderate disease** are:

$$IgM(N)\ (t) = 1.896 - 0.258e^{-12.053t} - 22.852(e^{-12.053t} - e^{-9.737t})$$

$$IgG(N)\ (t) = 0.166 + 3.297e^{-49.361t} - 8.193(e^{-49.361t} - e^{-8.194t})$$

$$IgG(S)\ (t) = 204.637 + 8155.607e^{-40.462t} - 8280.22e^{-40.462t}$$

Normalizing *t* according to the maximum length of follow-up (i.e., 360 for (N) antibodies, 270 for (S) antibody).

The adjusted equations for patients having a **severe to critical disease** are:

$$IgM(N)\ (t) = 1.371 + 5.811e^{-10.052t} - 13.781(e^{-10.052t} - e^{-5.074t})$$

$$IgG(N)\ (t) = 3.171 + 1.641e^{-8.447t} - 26.785(e^{-8.447t} - e^{-3.859t})$$

$$IgG(S)\ (t) = 138.898 + 12017.33e^{-26.121t} - 11464.45e^{-26.121t}$$

Normalizing *t* according to the maximum length of follow-up (i.e., 360 for (N) antibodies, 270 for (S) antibody).

The adjusted equations for **female** patients are:

$$IgM(N)\ (t) = 1.556 + 0.216e^{-12.144} - 18.896(e^{-12.144} - e^{-9.963})$$

$$IgG(N)\ (t) = 3.056 - 0.014e^{-8.347} - 25.508(e^{-8.347} - e^{-5.664})$$

$$IgG(S)\ (t) = 196.788 - 1285.91\ e^{-65.932} - 12974.63e^{-65.932}$$

Normalizing *t* according to the maximum length of follow-up (i.e., 360 for (N) antibodies, 270 for (S) antibody).

The adjusted equations for **male** patients are:

$$IgM(N)\ (t) = 1.566 + 0.378e^{-13.801t} - 21.612(e^{-13.801t} - e^{-10.754t})$$

$$IgG(N)\ (t) = 3.496 + 0.188e^{-10.454} - 36.547(e^{-10.454t} - e^{-7.314t})$$

$$IgG(S)\ (t) = 225.22 + 9440.56e^{-42.339t} - 9535.59e^{-42.339t}$$

Normalizing *t* according to the maximum length of follow-up (i.e., 360 for (N) antibodies, 270 for (S) antibody).

#### 4 REFERENCES

Pinheiro J, Bates D, DebRoy S, Sarkar D, R Core Team (2020). nlme: Linear and Nonlinear Mixed Effects Models. R package version 3.1-144. Available at: https://CRAN.R-project.org/package=nlme

**Table S1.** Number of available samples of each SARS-CoV-2 antibody per assessment timepoint.

| Timepoint (days since diagnosis) | Mean number of days of follow-up | IgM(N) samples | IgG(N) samples | IgG(S) samples |
|---|---|---|---|---|
| **0** | 9.56 ± 3.80 | 162 | 162 | 147 |
| **15** | 22.7 ± 4.38 | 214 | 214 | 190 |
| **30** | 43.1 ± 7.86 | 282 | 282 | 244 |
| **60** | 74.5 ± 7.60 | 352 | 352 | 289 |
| **90** | 119 ± 21.8 | 400 | 400 | 305 |
| **180** | 214 ± 22.0 | 275 | 275 | 239 |
| **270** | 310 ± 23.4 | 195 | 195 | 98 |
| **360** | 401 ± 21.4 | 109 | 109 | - |

*Notes:* Each timepoint represent a certain number of days since diagnosis.

**Table S2.** Description (N, %) of the main symptoms in participants according to disease severity and sex assigned at birth.

| Symptom, n (%) | Mild-moderate illness N = 367 (76.8) | | | | Severe-critical illness N = 39 (8.15) | | | | P-value |
|---|---|---|---|---|---|---|---|---|---|
| | Female N = 273 (74.4) | Male N = 94 (25.6) | P-value | Total | Female N = 19 (48.7) | Male N = 20 (51.3) | P-value | Total | |
| Headache | 190 (69.6) | 56 (59.6) | 0.10 | 246 (67.0) | 12 (63.2) | 11 (55.0) | 0.848 | 23 (59.0) | 0.41 |
| Diarrhea | 104 (38.1) | 32 (34.0) | 0.56 | 136 (37.1) | 9 (47.4) | 8 (40.0) | 0.888 | 17 (43.6) | 0.53 |
| Dyspnea | 61 (22.3) | 14 (14.9) | 0.16 | 75 (20.4) | 11 (57.9) | 8 (40.0) | 0.425 | 19 (48.7) | <0.001 |
| Fever | 140 (51.3) | 52 (55.3) | 0.58 | 192 (52.3) | 13 (68.4) | 19 (95.0) | 0.044 | 32 (82.1) | 0.001 |
| Cough | 152 (55.7) | 48 (51.1) | 0.51 | 200 (54.5) | 12 (63.2) | 9 (45.0) | 0.415 | 21 (53.8) | 1 |
| Anosmia | 146 (53.5) | 37 (39.4) | 0.03 | 183 (49.9) | 8 (42.1) | 7 (35.0) | 0.899 | 15 (38.5) | 0.24 |
| Arthralgias | 119 (43.6) | 26 (27.7) | 0.01 | 145 (39.5) | 11 (57.9) | 9 (45.0) | 0.628 | 20 (51.3) | 0.21 |
| Asthenia | 189 (69.2) | 50 (53.2) | 0.01 | 239 (65.1) | 16 (84.2) | 15 (75.0) | 0.695 | 31 (79.5) | 0.10 |
| Shivers | 95 (34.8) | 29 (30.9) | 0.57 | 124 (33.8) | 9 (47.4) | 8 (40.0) | 0.888 | 17 (43.6) | 0.30 |
| Chest pain | 40 (14.7) | 11 (11.7) | 0.59 | 51 (13.9) | 3 (15.8) | 3 (15.0) | 1.000 | 6 (15.4) | 0.99 |

| | | | | | | | | | |
|---|---|---|---|---|---|---|---|---|---|
| Epigastralgia | 42 (15.4) | 12 (12.8) | 0.65 | 54 (14.7) | 4 (21.1) | 5 (25.0) | 1.000 | 9 (23.1) | 0.26 |
| Discomfort | 160 (58.6) | 50 (53.2) | 0.43 | 210 (57.2) | 12 (63.2) | 14 (70.0) | 0.910 | 26 (66.7) | 0.33 |
| Myalgias | 143 (52.4) | 36 (38.3) | 0.03 | 179 (48.8) | 13 (68.4) | 14 (70.0) | 1.000 | 27 (69.2) | 0.02 |
| Nausea | 48 (17.6) | 12 (12.8) | 0.35 | 60 (16.3) | 4 (21.1) | 2 (10.0) | 0.407 | 6 (15.4) | 1 |
| Odinophagy | 74 (27.1) | 13 (13.8) | 0.01 | 87 (23.7) | 3 (15.8) | 1 (5.00) | 0.342 | 4 (10.3) | 0.09 |
| Congestion | 100 (36.6) | 25 (26.6) | 0.10 | 125 (34.1) | 2 (10.5) | 4 (20.0) | 0.661 | 6 (15.4) | 0.03 |
| Others | 70 (25.6) | 22 (23.4) | 0.77 | 92 (25.1) | 5 (26.3) | 1 (5.00) | 0.091 | 6 (15.4) | 0.25 |
| **Health service use, n (%)** | | | | | | | | | |
| Emergency room | 50 (18.3) | 20 (21.3) | 0.63 | 70 (19.1) | 16 (84.2) | 18 (90.0) | 0.661 | 34 (87.2) | <0.001 |
| Hospital | 0 (0) | 0 (0) | - | 0 (0.00) | 19 (100) | 20 (100) | . | 39 (100) | <0.001 |
| ICU | 0 (0) | 0 (0) | - | 0 (0.00) | 2 (10.5) | 4 (20.0) | 0.661 | 6 (15.4) | <0.001 |

*Notes:* Prevalence of symptoms stratified by sex was also study in each clinical condition. The p-value column shows the corresponding Chi-square test result. Asymptomatic participants were excluded from this analysis as they did not report any symptom. Prevalence of disease severity is calculated on the total of infected participants.

**Table S3.** Parameter estimation for SARS-CoV-2 antibodies (IgM(N), IgG(N), IgG(S)) NLME models.

| SARS-CoV-2 antibody | Model | Variance of random effects residuals | BIC | AIC | Parameter | Value (SE) | P-value |
|---|---|---|---|---|---|---|---|
| IgM (N) | Complete | 2.64 | 8645.24 | 8605.74 | b0 - Baseline value | 2.07 (0.17) | <0.001 |
| | | | | | b1 - Asymptotic value (i.e., antibody levels at t →∞) | 1.22 (0.14) | <0.001 |
| | | | | | k1 - Rise rate | 10.75 (0.52) | <0.001 |
| | | | | | k2 - Decay rate | 7.10 (0.62) | <0.001 |
| | IgM (N) ~ Clinical condition | 1.88 | 8398.88 | 8314.23 | b0 (Mild-moderate) | 1.64 (0.17) | <0.001 |
| | | | | | b0 (Asymptomatic) | 0.29 (0.31) | 0.34 |
| | | | | | b0 (Severe-critical) | 5.54 (0.89) | <0.001 |
| | | | | | b1 (Mild-moderate) | 1.90 (0.09) | <0.001 |
| | | | | | b1 (Asymptomatic) | 0.179 (0.23) | 0.44 |

| | | | | | | |
|---|---|---|---|---|---|---|
| | | | | b1 (Severe-critical) | -0.53 (0.30) | 0.08 |
| | | | | k1 (Mild-moderate) | 12.05 (0.37) | <0.001 |
| | | | | k1 (Asymptomatic) | 3.32 (1.15) | 0.004 |
| | | | | k1 (Severe-critical) | -2 (1.54) | 0.19 |
| | | | | k2 (Mild-moderate) | 9.74 (0.37) | <0.001 |
| | | | | k2 (Asymptomatic) | 4.88 (1.17) | <0.001 |
| | | | | k2 (Severe-critical) | -4.66 (0.81) | <0.001 |
| IgM (N) ~ Sex | 1.51 | 8610.53 | 8554.1 | b0 (Female) | 1.77 (0.14) | <0.001 |
| | | | | b0 (Male) | 0.17 (0.24) | 0.47 |
| | | | | b1 (Female) | 1.56 (0.07) | <0.001 |
| | | | | b1 (Male) | 0.01 (0.12) | 0.94 |
| | | | | k1 (Female) | 12.14 (0.34) | <0.001 |
| | | | | k1 (Male) | 1.66 (0.61) | 0.006 |
| | | | | k2 (Female) | 9.96 (0.34) | <0.001 |
| | | | | k2 (Male) | 0.79 (0.61) | 0.19 |

| | | | | | | | |
|---|---|---|---|---|---|---|---|
| | Complete | 4.71 | 10720.3 | 10680.8 | b0 - Baseline value | 4.17 (0.31) | <0.001 |
| | | | | | b1 - Asymptotic value (i.e., antibody levels at t →∞) | 2.24 (0.27) | <0.001 |
| | | | | | k1 - Rise rate | 9.72 (0.38) | <0.001 |
| | | | | | k2 - Decay rate | 5.62 (0.49) | <0.001 |
| **IgG (N)** | IgG (N) ~ Clinical condition | 4.56 | 10612.03 | 10527.38 | b0 (Mild-moderate) | 3.46 (0.42) | <0.001 |
| | | | | | b0 (Asymptomatic) | 0.13 (0.80) | 0.87 |
| | | | | | b0 (Severe-critical) | 7.44 (1.97) | <0.001 |
| | | | | | b1 (Mild-moderate) | 0.17 (0.03) | <0.001 |
| | | | | | b1 (Asymptomatic) | 1.79 (0.72) | 0.01 |
| | | | | | b1 (Severe-critical) | 3 (1) | 0.003 |
| | | | | | k1 (Mild-moderate) | 49.36 (8.79) | <0.001 |
| | | | | | k1 (Asymptomatic) | -39.50 (9.08) | <0.001 |
| | | | | | k1 (Severe-critical) | -40.91 (8.84) | <0.001 |
| | | | | | k2 (Mild-moderate) | 1.62 (0.14) | <0.001 |

| | | | | | | | |
|---|---|---|---|---|---|---|---|
| | | | | | k2 (Asymptomatic) | 6.20 (2.72) | 0.02 |
| | | | | | k2 (Severe-critical) | 2.24 (0.10) | 0.02 |
| | IgG (N) ~ Sex | 4.84 | 12585.17 | 12528.74 | b0 (Female) | 3.04 (0.43) | <0.001 |
| | | | | | b0 (Male) | 0.64 (0.77) | 0.4 |
| | | | | | b1 (Female) | 3.06 (0.42) | <0.001 |
| | | | | | b1 (Male) | 0.44 (0.66) | 0.5 |
| | | | | | k1 (Female) | 8.35 (0.84) | <0.001 |
| | | | | | k1 (Male) | 2.11 (1.37) | 0.13 |
| | | | | | k2 (Female) | 5.66 (1.10) | <0.001 |
| | | | | | k2 (Male) | 1.65 (1.66) | 0.32 |
| **IgG (S)** | Complete | 213.83 | 26009.03 | 25975.6 | b0 - Baseline value | 8202.59 (4221.75) | 0.05 |
| | | | | | b1 - Asymptotic value (i.e., antibody levels at t $\rightarrow\infty$) | 218.62 (7.91) | <0.001 |
| | | | | | k1 - Rise rate | 37.01 (19.77) | 0.06 |

| | | | | | | |
|---|---|---|---|---|---|---|
| IgG (S) ~ Clinical condition | 204.85 | 25887.21 | 25820.36 | b0 (Mild-moderate) | 8360.24 (4493.10) | 0.06 |
| | | | | b0 (Asymptomatic) | -9386.21 (39058.58) | 0.81 |
| | | | | b0 (Severe-critical) | 4095.98 (6944.90) | 0.56 |
| | | | | b1 (Mild-moderate) | 204.64 (8.55) | <0.001 |
| | | | | b1 (Asymptomatic) | -67.50 (20.66) | 0.001 |
| | | | | b1 (Severe-critical) | 234.26 (26.26) | <0.001 |
| | | | | k1 (Mild-moderate) | 40.46 (22.50) | 0.07 |
| | | | | k1 (Asymptomatic) | -48.94 (284.18) | 0.86 |
| | | | | k1 (Severe-critical) | -14.34 (25.29) | 0.57 |
| | 207.07 | 28243.93 | 28198.78 | b0 (Female) | 13054.70 (23205.04) | 0.57 |

| | | | |
|---|---|---|---|
| IgG (S) ~ Sex | b0 (Male) | -3388.92 (26483.37) | 0.9 |
| | b1 (Female) | 196.79 (7.37) | <0.001 |
| | b1 (Male) | 28.43 (13.48) | 0.04 |
| | k1 (Female) | 65.93 (118.85) | 0.58 |
| | k1 (Male) | -23.59 (132.06) | 0.86 |

AIC: Akaike information criterion; BIC: Bayesian information criterion; SE: standard error

*Notes:* Three different models were fitted for each antibody: the first one was general, the second one was stratified by clinical condition, and the third one was stratified by sex assigned at birth. The reference categories were mild-moderate and female. The parameters of the other categories are expressed with reference to these categories. The fitted equations of these models can be found in section 2.3 of the Supplementary Methods.

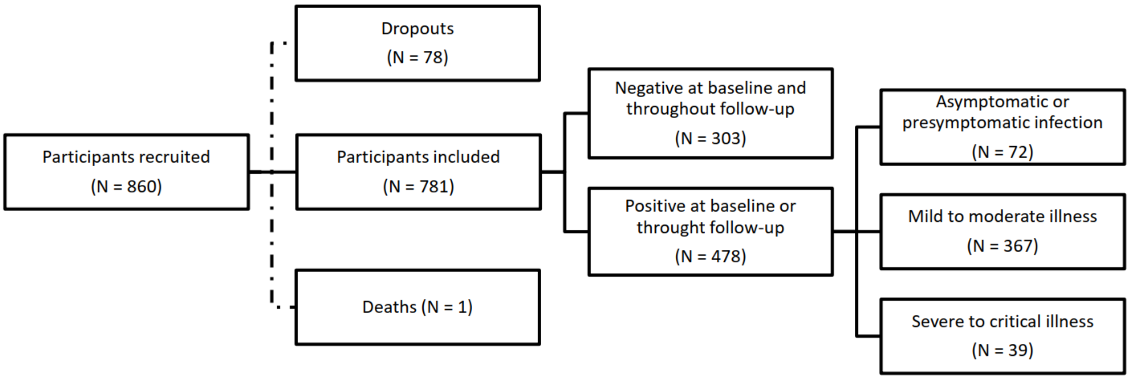

**Figure S1**. Flow chart of the ProHEpiC-19 study participants, including the recruitment procedure and the type of relationship with Sars-CoV-2.
